# SARS-CoV-2 titers in wastewater foreshadow dynamics and clinical presentation of new COVID-19 cases

**DOI:** 10.1101/2020.06.15.20117747

**Authors:** Fuqing Wu, Amy Xiao, Jianbo Zhang, Katya Moniz, Noriko Endo, Federica Armas, Richard Bonneau, Megan A Brown, Mary Bushman, Peter R Chai, Claire Duvallet, Timothy B Erickson, Katelyn Foppe, Newsha Ghaeli, Xiaoqiong Gu, William P Hanage, Katherine H Huang, Wei Lin Lee, Mariana Matus, Kyle A McElroy, Jonathan Nagler, Steven F Rhode, Mauricio Santillana, Joshua A Tucker, Stefan Wuertz, Shijie Zhao, Janelle Thompson, Eric J Alm

## Abstract

Current estimates of COVID-19 prevalence are largely based on symptomatic, clinically diagnosed cases. The existence of a large number of undiagnosed infections hampers population-wide investigation of viral circulation. Here, we use longitudinal wastewater analysis to track SARS-CoV-2 dynamics in wastewater at a major urban wastewater treatment facility in Massachusetts, between early January and May 2020. SARS-CoV-2 was first detected in wastewater on March 3. Viral titers in wastewater increased exponentially from mid-March to mid-April, after which they began to decline. Viral titers in wastewater correlated with clinically diagnosed new COVID-19 cases, with the trends appearing 4-10 days earlier in wastewater than in clinical data. We inferred viral shedding dynamics by modeling wastewater viral titers as a convolution of back-dated new clinical cases with the viral shedding function of an individual. The inferred viral shedding function showed an early peak, likely before symptom onset and clinical diagnosis, consistent with emerging clinical and experimental evidence. Finally, we found that wastewater viral titers at the neighborhood level correlate better with demographic variables than with population size. This work suggests that longitudinal wastewater analysis can be used to identify trends in disease transmission in advance of clinical case reporting, and may shed light on infection characteristics that are difficult to capture in clinical investigations, such as early viral shedding dynamics.

## Introduction

The ongoing coronavirus disease (COVID-19) pandemic, caused by severe acute respiratory syndrome coronavirus 2 (SARS-CoV-2), has quickly become a global health crisis, with over 2 million confirmed cases in the US and 7.4 million worldwide as of June 11, 2020 (1). Due to limited diagnostic capacity in many countries and high rates of asymptomatic individuals (2,3), these numbers are considered to be underestimates of the true prevalence of infection (4,5).

Wastewater surveillance offers a complementary approach to clinical disease surveillance (6,7) as it aggregates health information at the population level. In addition, wastewater surveillance provides an unbiased sample of the infected population, including asymptomatic and pre-symptomatic individuals, those who are symptomatic but have not yet been clinically confirmed, and individuals who may have the disease but do not seek healthcare. Early work has shown that wastewater surveillance could detect SARS-CoV-2 before it became widespread in a population (8), and that viral levels in wastewater largely paralleled local increases in clinical cases of COVID-19 in France (9), Spain (10), Turkey (11), and Israel (12). These studies highlight the potential of wastewater surveillance to provide early warning of emerging outbreaks – as well as the need for improved modeling and quantitation to understand the relationship between viral concentrations in wastewater and active infections in the population.

The Massachusetts wastewater treatment facility where we obtained samples has two major influent streams, which we will refer to as the “northern” and “southern” influents. Together the two catchments represent approximately 2.25 million sewered individuals in Middlesex, Norfolk, and Suffolk counties, primarily in urban and suburban neighborhoods. We previously reported the detection of SARS-CoV-2 in wastewater from this facility, and showed that viral titers were significantly higher than would be expected based on clinical cases alone (13). Here, we use longitudinal sampling from January to mid May 2020 (116 samples in total) at the same wastewater treatment facility to identify the virus’s first appearance and spread, to infer viral shedding dynamics early in the course of infection, and to investigate the relationship between wastewater SARS-CoV-2 titers at different spatial scales, clinically reported COVID-19 cases, and statewide public health interventions.

## Results and Discussion

We first quantified SARS-CoV-2 titers in 116 longitudinal wastewater samples from the wastewater treatment facility’s northern and southern influents, from January to May 2020. Samples from March 3 to May 20 were processed a second time in late May, in a single batch for each catchment, to complement initial quantification, which had been performed in chronological groups as samples were received (see Methods).

### Longitudinal wastewater sampling captures the emergence and spread of SARS-CoV-2 in the population

Viral titers in wastewater followed a trend similar to new clinical cases over the sampled time period (Figure 1A). SARS-CoV-2 was not detected in either influent stream in January or February samples, and was first detected on March 3 (northern influent) and March 10 (southern influent), at a concentration of ≤15 copies per ml of wastewater (Figure 1A and Figure S1). Only two clinically confirmed COVID-19 cases had been reported in Massachusetts as of March 3, indicating viral detection in wastewater in the early stage of the local outbreak. SARS-CoV-2 titers remained low (< 90 copies per ml) in both influents until March 15, after which they increased exponentially. Viral titers appeared to rise in wastewater in advance of clinical cases. SARS-CoV-2 titers in wastewater began trending downward about a month later (April 13), while the decline in new clinical cases peaked later, on April 24. Together, these qualitative trends suggested that wastewater viral titers might reflect disease incidence in advance of clinical reporting.

**Figure 1.**
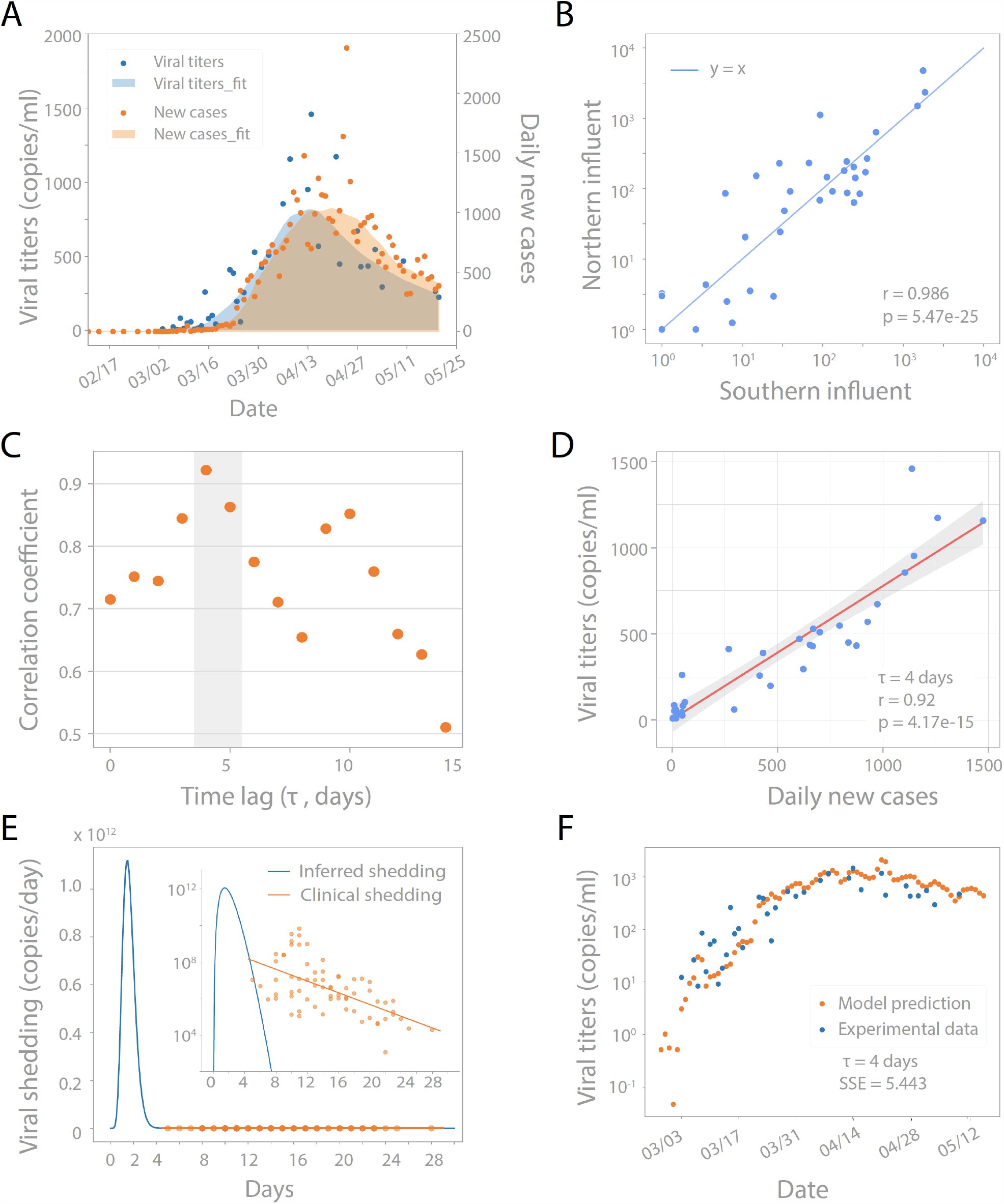
SARS-CoV-2 titers in wastewater correlate with new clinical cases, with a temporal offset. (A) Viral titers in wastewater samples from March 3 to May 20 (blue dots) and new clinical cases from February 12 to May 20 in Norfolk, Suffolk, and Middlesex counties served by the wastewater treatment plant (orange dots). LOWESS smoothing was applied to show the trends for viral titers (shaded blue) and daily new cases (shaded orange). (B) Unsmoothed viral titers in the northern and southern influents are highly correlated and have similar magnitudes when run in chronological batches using Method I (see Methods). Blue line represents y=x. Pearson’s r = 0.986, p = 5.47e-25. (C) Linear correlation between unsmoothed viral titers in wastewater and unsmoothed daily new cases with different time lags from 0 to 14 days. Pearson correlation coefficient is highest with a 4 day time lag. (D) Viral titers correlate with daily new cases with a 4 day time lag. Red solid line is the linear regression fitting. Grey area: 95% confidence interval from standard error of the fitting. Pearson’s r = 0.92, p = 4.17e-15. (E-F) Modeling wastewater titers as a convolution of new cases per day and virus shedding per day. (E) Beta function with optimal shape and scaling parameters (*α, β, c*) representing an individual’s virus shedding pattern *s*(*t*) on a linear scale and log scale (inset). This optimal shedding function minimizes the sum of squared errors between model predictions and observed wastewater data. The raw beta function is multiplied by an estimate of wastewater volume to report total copies shed per individual per day. Clinically reported values are added in orange for reference, with linear regression fit (18). (F) Viral titers predicted by convolution model compared to viral titers observed in wastewater. Model predictions are the convolution of new cases per day *I*(*t*) and optimal shedding function s(*t*) from (E). SSE: sum of squared errors. *τ*: clinical data time lag.

SARS-CoV-2 was consistently detected at comparable levels, and demonstrated similar dynamics, in the northern and southern influents (Figure 1B), which were combined for further analyses.

### Wastewater data correlates best with clinically confirmed new cases, with a temporal offset

Since SARS-CoV-2 was first detected in wastewater when only two cases were clinically confirmed, we hypothesized that the wastewater signal included a significant undiagnosed, COVID-19 positive population. This difference between wastewater and clinical data could be due to underdiagnosis of asymptomatic or mildly symptomatic cases, limitations in clinical testing capacity, or a time delay between viral shedding and the onset of respiratory and other symptoms.

We tested the correlation of viral titers with new clinical cases and cumulative cases, allowing for a variable time lag (Figure 1C/D and Figure S2). Higher correlations were seen when comparing new clinical cases back-dated by 4-10 days than for shorter or longer temporal offsets, and the maximum agreement between the two time series was observed for a time offset of 4 days (Pearson’s r = 0.92, p = 4.17e-15, Figure 1C/D). Similar results were found when each catchment was considered individually (Figure S3). This time lag between the wastewater signal and clinically reported cases is consistent with the typical 4-5 day incubation period from SARS-CoV-2 infection to symptom onset (14-16). Moreover, a short time lag in an exponentially growing process explains large differences in the total number of cases (*e*.*g*. 2 cases as of March 3, compared to 108 cases as of March 12). Thus, wastewater surveillance could potentially be used to predict trends in new COVID-19 cases.

### An inferred viral shedding function suggests an early burst of high viral shedding

The high correlation between wastewater viral titers and daily reports of new clinically confirmed COVID-19 cases (Figure 1D) – combined with the time lag between the wastewater signal and clinical data – suggests that newly infected individuals contribute significant viral loads to the wastewater and that most of this shedding occurs early in infection, prior to the individual seeking healthcare and being tested. As a result, viral dynamics in wastewater could shed light on early shedding, which would be challenging to capture clinically if it precedes patient presentation to the clinic.

We attempted to infer average individual viral shedding dynamics by comparing our longitudinal wastewater data with daily reports of new clinical cases. We modeled the wastewater data *W*(*t*) as a convolution of new clinical cases *I*(*t*) and a function *s*(*t*) describing viral shedding (whether from fecal, urine, or other unknown sources) into wastewater by each infected individual. To obtain *I*(*t*), clinical data was back-dated to account for the time lag with wastewater data. We optimized the parameters of the shedding function to minimize the error between the convolutional model’s prediction and the observed wastewater titers.

Modeling revealed a short burst of viral shedding lasting 3-4 days. This feature of the modeling was robust to different time lags (0-10 days), as well as the distribution used to fit the shedding function (both beta and gamma distributions were investigated, Figure S4). This short burst of shedding was also seen after modeling each catchment individually (Figure S5-6). Using a 4-day time lag, which gave the best correlation between wastewater and clinical data (Figure 1C-D), we found that the individual shedding function peaked around day 2 and spanned approximately 3-4 days (Figure 1E). Peak shedding was two orders of magnitude greater than the highest clinically reported abundance of SARS-CoV-2 in stool, and generally several orders of magnitude greater than typical values, which are measured after symptom onset and clinical testing (Figure 1E, Ref. 17, Ref.18).

The inferred viral shedding function allows us to estimate the viral titers in wastewater based on reported clinical cases. Figure 1F shows new clinical cases, back-dated by 4 days, convolved with the inferred shedding function to generate estimated viral titers in wastewater. These data fit well with observed viral titers. It should be noted, however, that a good fit could be obtained even if a large fraction of cases are underreported. For example, if only half of cases are reported, then the total amount of shedding per individual would be inferred to be twice the actual value, even though the fit to the model would be equally good.

Together, our data suggest that SARS-CoV-2 levels in wastewater may be largely driven by a burst of shedding that occurs early in infection – likely before the onset of respiratory symptoms commonly used as testing criteria. Although the early peak of viral shedding in our model does not directly reflect patients’ reported clinical courses, it is consistent with reports of abdominal pain, nausea, and diarrhea preceding onset of respiratory symptoms in COVID-19 patients (19-21), suggesting that individuals may shed SARS-CoV-2 virus early in this process. Our data is also consistent with recent findings in patients’ oropharyngeal swab samples (peak shedding at 0.7 days before onset of respiratory symptoms, Ref. 22), and supported by observations in a transgenic mouse model with humanized ACE2 receptors (peak titers in intestines at 1 day post infection, Ref. 23).

We hypothesize that viral shedding is characterized by an early burst, inferred by our model, followed by a period of prolonged low-level shedding as reported in the literature (17, 18, 24). In several clinical studies, the earliest stool samples collected have been positive for viral RNA (day 3 post symptom onset, Ref 18; and day 0 of hospital admission, Ref. 24), suggesting fecal shedding may start before individuals seek medical care. Given the orders of magnitude difference between our inferred peak of shedding (10^12 copies per day) and the median clinically reported shedding (10^6.11 copies per day, assuming one 200 g stool per day, Ref. 18), persistent low-level shedding for 1-2 months is unlikely to interfere with the sensitivity of wastewater surveillance to detect new infections above the background of previous cases. Our model, which suggests peak viral shedding at levels up to 1.1 x 10^12 viral copies per day, also helps to explain our previously reported findings, that SARS-CoV-2 levels in wastewater are significantly higher than would be expected based on clinical case numbers and observations of viral shedding in patient stool (13).

### Wastewater SARS-CoV-2 titers in the context of behavior and interventions

SARS-CoV-2 levels in wastewater began to increase exponentially after March 15, coinciding with a peak in reports to the US Centers for Disease Control and Prevention of “influenza-like illnesses” (ILI) in Massachusetts that were not caused by influenza (Figure 2, panel 3) – and which showed early dynamics similar to SARS-CoV-2 levels in wastewater, suggesting these ILIs may have been undiagnosed COVID-19 (4, 25, 26). Wastewater titers of SARS-CoV-2 began to drop in mid-April (Figure 1A), roughly one month after the state of emergency was declared (March 10, Figure 2) and the statewide school closure (March 17; Figure 2), and approximately three weeks after the Massachusetts stay-at-home advisory went into effect (March 24, Figure 2) (27-29). Public transport and cellular mobility data indicate that public movement began to decrease significantly ahead of the stay-at-home advisory, starting soon after the state of emergency was declared and the first school closures (30, 31). The time between infection and forward transmission is approximately 4-6 days (22, 32), suggesting that there were likely 4-5 additional cycles of infection after the implementation of the stay-at-home advisory before viral titers began to decline in wastewater.

**Figure 2.**
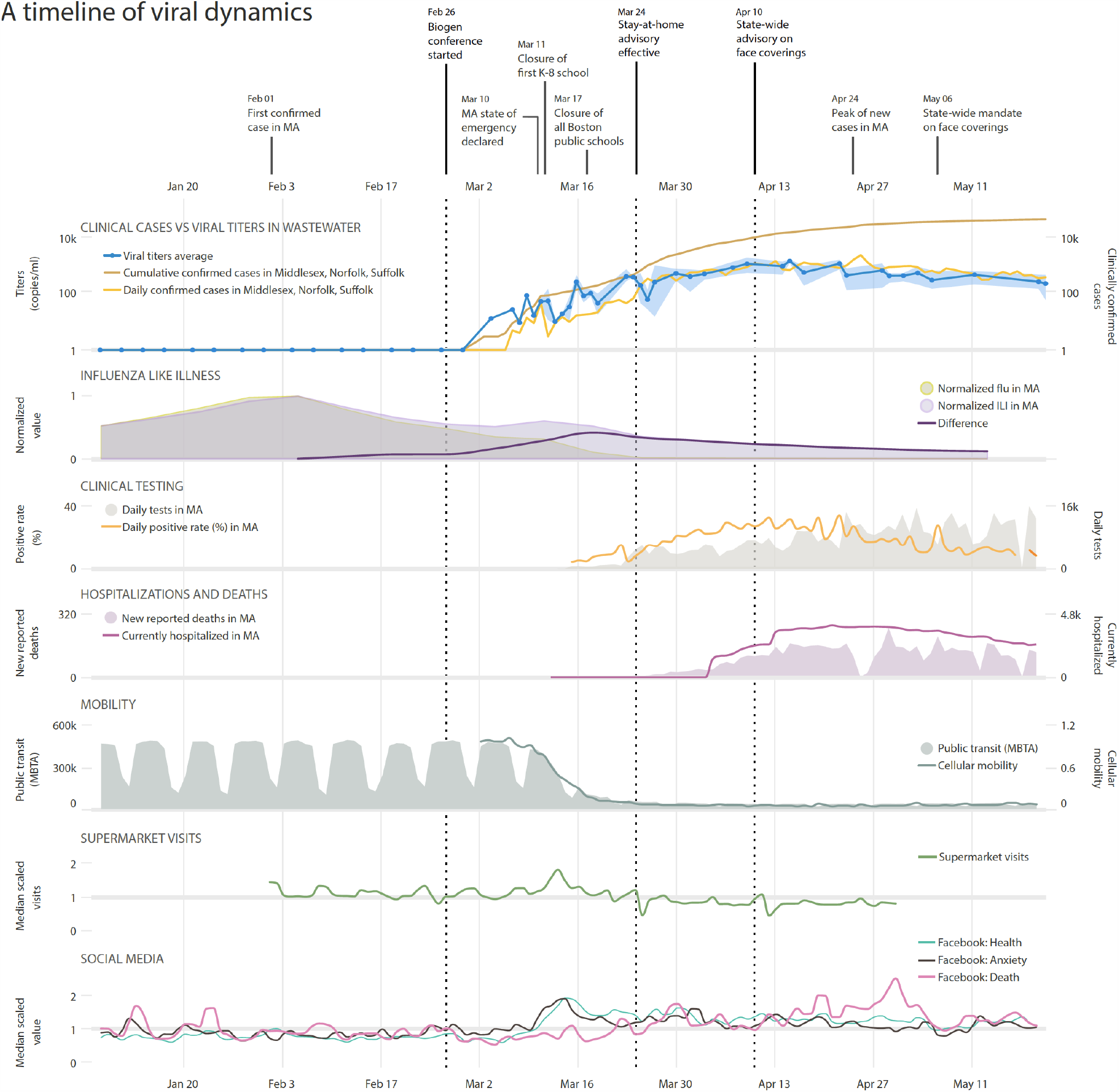
A timeline of viral dynamics in the context of key events and clinical/behavioral data. Trends are plotted in the same time frame, from January 8 to May 20. Row 1: Timeline of COVID-19 pandemic and important events in MA. Clinical Cases vs Viral Titers in Wastewater: Viral titers in wastewater (blue line along the primary y-axis, shaded area represents the minimum/maximum of normalized and averaged values), daily (orange line) and cumulative (brown line) confirmed cases along the secondary y-axis. Influenza Like Illness: Visits for influenza-like illness (ILI, purple shading) and confirmed flu cases (light green shading), and the difference between the two after normalization (purple line), which shows a peak of non-flu ILI at March 18. Clinical Testing: Daily SARS-CoV-2 tests and positive rates in MA. Hospitalizations and Deaths: New reported COVID-19 related deaths and hospitalizations in MA. Mobility: Public transit and cellular mobility data. Supermarket Visits: Supermarket visits in MA (normalized by the median value). Social Media: Facebook posts with terms expressing “Health”, “Anxiety”, and “Death”. Dashed lines in all the panels represent the date of the Biogen conference, the stay-at-home advisory in MA, and the state-wide face covering advisory.

What could explain the approximately month-long lag between decreased public mobility and the drop in population-wide viral levels as measured in wastewater? Several factors likely contributed. First, despite efforts to limit human-to-human contact, disease transmission could have remained high among especially at-risk populations such as essential workers and their families. Second, individuals infected before the stay-at-home order could have infected multiple family members or roommates in subsequent days, especially in areas with high numbers of inhabitants per household. Finally, despite the stay-at-home order, human-to-human interactions may still have occurred at a significant rate outside the household, in essential venues such as supermarkets and pharmacies. These early interactions may have been more likely to result in disease transmission because the Massachusetts advisory recommending face coverings in public was not announced until April 10 (33), and the state order requiring face coverings did not go into effect until May 6 (34). Social media indices of public anxiety showed a pronounced rise in anxiety around the time of the declaration of the state of emergency (35), followed shortly thereafter by a peak in Massachusetts supermarket traffic on March 13 (Figure 2, panel 7-8; Ref. 36). This concentration of people into supermarkets – without masks, and before the institution of supermarket policies to limit the number of shoppers – might have contributed to the sharp rise in viral titers in wastewater (Figure 1A) that began in the following days.

### Wastewater viral titers at the neighborhood level correlate better with demographic variables than with population size

Whatever the cause of the additional infection cycles after the public began to stay at home in mid-March, was the exponential rise in viral titers uniformly distributed across the catchments we sampled? The wastewater treatment facility’s northern and southern influents showed similar SARS-CoV-2 levels and dynamics (Figure 1B) – suggesting that disease incidence (new cases) and/or prevalence (total cases) were similar in both catchments. We hypothesized that similar viral levels in the two influents were due to a relatively uniform spatial distribution of disease burden across both catchments, which represent similar population sizes. An equal distribution of disease incidence across the catchment would mean that the likelihood of seeing a positive SARS-CoV-2 signal should increase with the size of the catchment according to Poisson statistics (Figure S8). Thus, comparing presence/absence of disease to a binomial (Poisson) model of infection would enable more accurate estimates of disease incidence, and would allow us to calibrate the observed viral titers to the true number of underlying cases.

To test this approach to calibrating viral titers to the number of cases, we sampled 11 urban neighborhoods within the wastewater treatment facility’s catchment (see Methods), representing populations ranging from ∼4,000 to ∼40,000 individuals. The observed viral titers deviated strongly from that expected under a Poisson process (Fig. 3A and Figure S8B). Many small catchments were found to be positive while larger catchments were negative, suggesting significant heterogeneity in disease levels between different neighborhoods. It should be noted that while some neighborhood catchments did not have detectable viral titers in sewage at the time they were sampled, new clinical cases were reported during the week of sampling.

**Figure 3.**
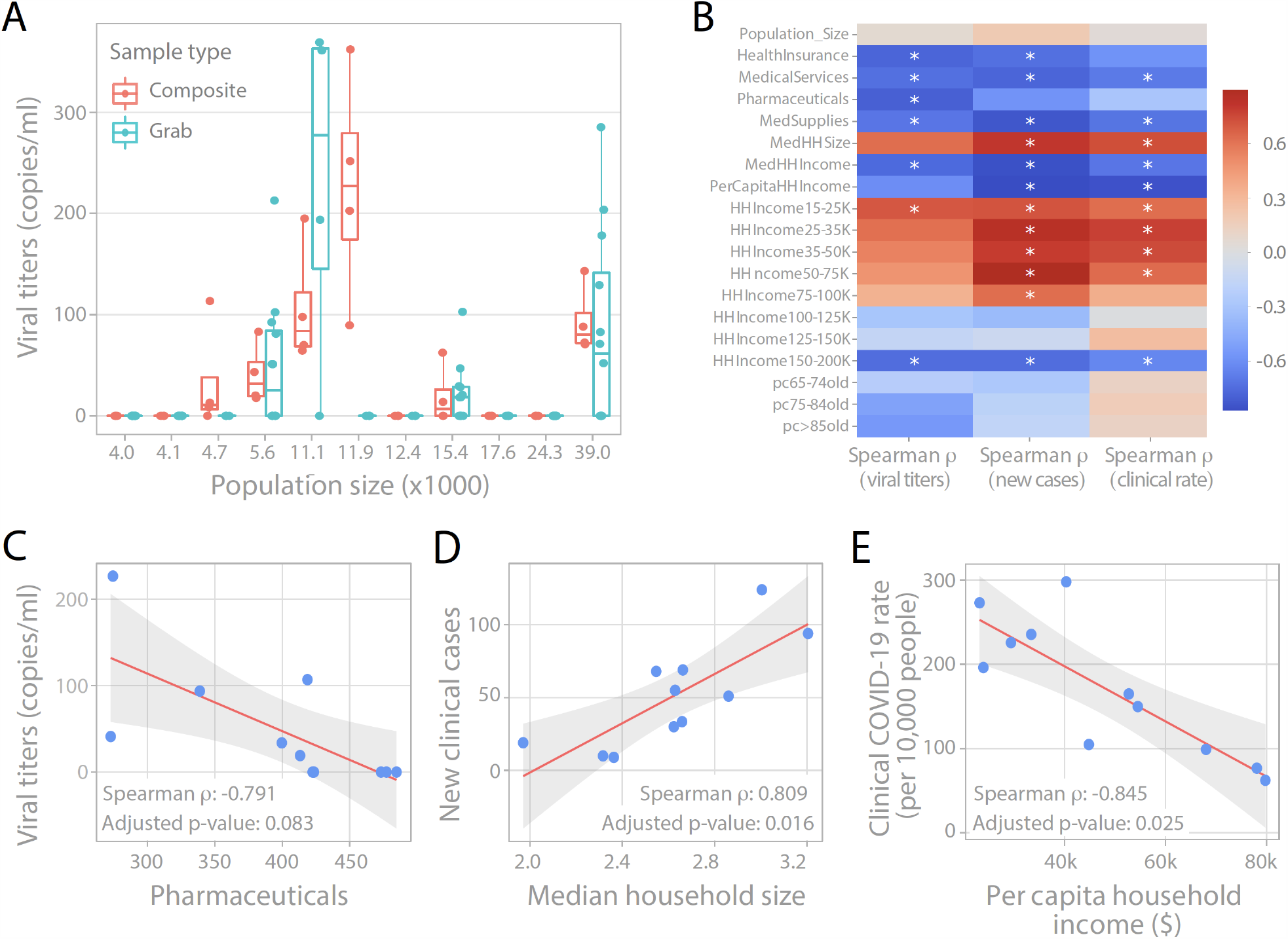
Wastewater viral titers at the neighborhood level correlate better with demographic variables than with population size. (A) Viral titers in the composite or grab wastewater samples of 11 neighborhoods with varying population sizes in a Massachusetts city, processed with Method I. (B) Correlation between wastewater viral titers (left), or new clinical cases (middle), or cumulative clinical case rate (right) and demographic data of the 11 catchments. Colormap shows Spearman’s rank correlation coefficient. Asterisk denotes significant p-value at FDR < 0.1. Full data is shown in Figure S9. HealthInsurance, MedicalServices, Pharmaceuticals, MedSupplies: average household expenditure on health insurance, medical services, pharmaceuticals, and medical supplies. MedHHSize: Median household size. MedHHIncome, PerCapitaHHIncome: Median and Per capita household income. HHIncome15-25K: percentage of population with household income within the range 15-25K per year. pc65-74old: percentage of individuals aged 65-74 years. (C) Correlation between wastewater viral titers and average household expenditure for pharmaceuticals. (D) Correlation between new COVID-19 cases during the week of sampling and median household size. (E) Correlation between cumulative clinically reported COVID-19 rate per 10,000 people and per capita household income. Red solid line is the linear regression fitting. Grey area: 95% confidence interval from standard error of the fitting.

To understand which factors were associated with this microgeographic heterogeneity, we investigated 38 different demographic variables. We found that viral levels in wastewater did not correlate strongly with population size, as would be expected under the null model of equal incidence (Figure 3B). Instead, wastewater titers correlated most strongly with average household healthcare spending and household income (left panel in Figure 3B; and Figure 3C). The associations between wastewater titers and demographic variables were broadly the same as those from clinical observations. An analysis of new clinical case reports for the neighborhoods containing the catchments during the week of sampling showed that new clinical cases correlated with household income, household size, age, healthcare spending, and race/ ethnicity (middle panel in Figure 3B and Figure S9; and Figure 3D). We note that many of these variables are correlated with each other. Similar trends were also found for cumulative clinical case rates (right panel in Figure 3B and Figure S9; and Figure 3E). Thus, wastewater surveillance at the neighborhood level reflects heterogeneity in disease incidence and prevalence in specific sub-populations, and is consistent with previous reports that the COVID-19 pandemic is disproportionately affecting neighborhoods with more socioeconomically disadvantaged populations (37-39). As a next step, we are continuing our neighborhood-level sampling to investigate viral loads in different communities.

### Uncertainties and future directions of wastewater surveillance

Although wastewater viral titers should be relatively unbiased, relatively high day-to-day variation in our data suggests that they are noisy even when disease incidence is high. On the other hand, clinical data may be less noisy but more prone to systematic bias due to constraints such as limited clinical testing capacity. Together, these independent data streams from clinical- and wastewater-based surveillance can provide a more complete picture of viral dynamics, and a new opportunity to explore the relationship between COVID-19 dynamics and concomitant public health interventions and behavioral changes. In this study, we infer a viral shedding function that can help to connect these two data streams in a quantitative fashion to allow for more accurate epidemiological modeling – and ultimately, more informed decision-making about how and when to intervene.

Several knowledge gaps introduce uncertainty to our modelling approaches. First, viral titers in wastewater are highly dependent on individual shedding rates, viral stability in wastewater, and flow rates (dilution) of the influent. While we have noted that normalization with the human fecal indicator PMMoV (see Methods) helps reduce noise in our datasets that is due to sampling time and flow, significant uncertainties remain regarding the consistency of viral shedding among COVID-19 patients (17, 24). Second, the viral shedding profile inferred by our modelling approach relies on reported clinical new cases per day over the course of the pandemic, and is thus subject to the limitations of clinical testing capacity (Figure 2). This early lack of testing is especially evident given that SARS-CoV-2 was detectable in wastewater when there were only 2 confirmed cases in the catchment (Figure 1A). Close examination of the trends of influenza like illnesses likely not attributable to influenza, as captured by ILI Net (26) suggest that people with ILI symptoms caused by COVID-19 may have been recorded by the CDC surveillance systems as early as the end of February in Massachusetts (Purple curve in the panel 2 of Figure 2; Ref. 4). We recognize that inadequate testing capacity, changing clinical criteria for testing eligibility, and the existence of populations who do not seek medical care could all influence the observed clinical dynamics – and in turn, the shape of the inferred shedding function. However, the alignment of emerging clinical and experimental evidence of early viral shedding with the shedding function inferred from wastewater analysis in this study lends credence to wastewater surveillance as a means to capture signatures from early disease dynamics.

This work demonstrates the power of longitudinal wastewater surveillance in tracking the emergence and spread of an infectious disease in a population, in advance of clinical case reporting -- as well as its potential to shed light on infection characteristics that may be challenging to capture clinically. It also highlights several key questions for future investigation. First, is SARS-CoV-2 excreted in urine as well as feces during the earliest stages of infection, before individuals seek testing? Occasional reports of SARS-CoV-2 detection (40, 41), including infectious virus (41), in patient urine suggest it could be possible. If it is consistently excreted in urine as well as stool, wastewater surveillance could be extended to include non-residential areas, allowing for disease monitoring at workplaces and schools as regions continue to re-open. Second, to what degree can our collective insights from SARS-CoV-2 wastewater surveillance be applied to other infectious diseases? Whether or not they generalize will depend on how excreted pathogen (or other infection biomarkers) signals relate to the course of each individual disease. This relationship may affect the sensitivity and quantitativeness of wastewater surveillance efforts, as well as determine whether wastewater signals correlate best with new cases or cumulative cases, if at all. Finally, this work highlights the importance of addressing batch effects in wastewater-based disease monitoring, where rapid availability of data is critical to enable real-time public health responses to disease dynamics. As we collectively explore the limitations and future directions of wastewater surveillance, its application in communities at this time can provide important guidelines for responding to the current or future public health crisis.

## Methods

### Sample collection and viral inactivation

24-hour composite samples of raw sewage were collected from the Deer Island Wastewater Treatment Plant (including the northern and southern influents) in Massachusetts. The samples were received in chronological groups, and then pasteurized at 60°C for 1 - 1.5 hours to inactivate the virus. Raw sewage was then vacuum filtered through a 0.2 µm membrane (Millipore Sigma) to remove bacterial cells and debris. Filtrate was sealed and stored at 4°C.

For the upstream catchment sampling, 24 hour composite samples and grab samples were collected from municipal sewer lines in 11 neighborhoods in a Massachusetts city from May 11-14, 2020. These 11 neighborhoods are spatially spread across the city, with population sizes ranging from 4,000 to 39,000 individuals. One 24 hour composite sample was taken at each of the 11 neighborhoods, drawing wastewater samples every 3 minutes over 24 hours. Grab samples were collected right before composite sampling started. Three grab samples were taken at 10 min intervals at six sites. Single grab samples were taken at five sites. Upon retrieval, sewage samples were transported on ice. Sewage samples were pasteurized and filtered as described for samples from the wastewater treatment facility (above), on the same day as collection, and processed using ‘Method I’, below.

### Viral precipitation, RNA extraction, reverse transcription and quantitative PCR (Method I)

The filtrate (40 mL) was mixed with 4 g of Polyethylene glycol 8000 (10% w/v, Millipore Sigma) and 0.9 g NaCl (0.3 M, Millipore Sigma) and centrifuged at 12,000 g at 4 °C for 2 hours (or overnight spinning with 3,200 g) to precipitate the viral particles. The viral pellet was then resuspended in 1.5 mL Trizol reagent (Cat# 15596026, Thermo Fisher Scientific) for RNA extraction. cDNA was synthesized by reverse transcription (RT) based on the manufacturer’s protocol (M0368, New England Biosciences). Briefly, 10 µl of RNA was mixed with random hexamers and then incubated at 70°C for 5 min and 4°C for 3 min. After that, the RNA-hexamer mixture was mixed with 5X ProtoScript II buffer (5 µl), 0.1 M DTT (2.5 µl), ProtoScript II Reverse Transcriptase (200 U/µl. 1.25 µl), 10 mM dNTP (1.25 µl), RNase Inhibitor (40 U/µl, 0.5 µl), and Nuclease-free water (2.5 µl) to a total volume of 25 µl. The mixture was then incubated at 42°C for 1 hour and inactivated at 65°C for 20 minutes.

The quantitative PCR (qPCR) was performed with TaqMan assay techniques. Briefly, the TaqMan® Fast Advanced Master Mix (4444557, ThermoFisher Scientific) was mixed with the primers and water, and then distributed into the 96-well PCR plate. 2 µl of cDNA template from RT was added into the plate, which was sealed with adhesive PCR plate seals (AB0558, ThermoFisher Scientific). The qPCR reaction was carried out for 48 cycles using Bio-Rad CFX96 Real-Time PCR Detection System based on the following program: polymerase activation (95 °C for 2 min), PCR (48 cycles, denature at 95°C for 1 s, and anneal/extend at 55°C for 30 s). The primers (N1 and N2) and DNA standards of SARS-CoV-2 nucleocapsid gene were used to quantify the titers of SARS-CoV-2 (42). Two or three replicates were performed for each primer, and mean values were reported.

Concentrations of fecal materials in wastewater are subject to wide fluctuations in daily sewage flow rates at the wastewater treatment plant, especially during this dynamic winter-spring season. To correct those variations, we used pepper mild mottle virus (PMMoV), a positive-strand RNA virus prevalent in human feces (43–45), as an internal reference for quantification. First, we used qPCR to evaluate the PMMoV concentrations across 108 sewage samples in our time-series (Southern and Northern influents) with following primers: (Forward: *5’-GAGTGGTTTGACCTTAACGTTGA-3’*; Reverse: *5’-TTGTCGGTTGCAATGCAAGT-3’*; Probe: *5’-FAM/CCTACCGAAGCAAATG/BHQ-3’* (36). Results showed that PMMoV is detectable and relatively stable between daily samples, with a standard deviation of 1.789 Ct value in the 108 tested samples (Figure S7). To adjust the SARS-CoV-2 viral titers for each sample, we first calculated the deviation of its PMMoV copies from the median of PMMoV copies in all 108 samples, i.e., deviation factor = 1^(*k* * (sample *Ct* - median *Ct*)), where *k* is the slope of the standard curve and equals to −0.2991 (amplification efficiency is 99.11% for this primer set) based on our test. We then divided the SARS-CoV-2 viral titers by this deviation. Our results showed that the data after adjustment is much less noisy and has a good correlation with the clinical trend. Thus, this data supports that PMMoV is a good internal reference for standardizing detection of RNA virus in sewage samples, and we use it to correct for sampling fluctuations and experimental batch effects.

We processed 108 longitudinal raw wastewater samples from northern and southern influents using this method (‘Method I’), in batches as they were received in the lab, from January 8 to May 5, 2020 (Figure S1).

### Viral precipitation, RNA extraction and RT-qPCR (Method II)

For Method I, raw wastewater samples were processed as they were received, in chronological groups -- providing near-real time information on viral titers in sewage, but raising the possibility of batch effects in virus precipitation or RNA extraction. Therefore, we reprocessed 60 samples from March 3 to May 20 (including 8 new samples from May) with a new method (‘Method II’) to allow all samples from a single influent to be processed together as a single batch. 15 ml of filtrate were first concentrated with 10 kDa Amicon Ultra Centrifugal Filter (Sigma, Cat# UFC9010) to 150 ∼ 200 ul, which is further lysed with 600 ul AVL buffer (Qiagen, Cat# 19073) for RNA extraction (Qiagen RNeasy kit, Cat# 74182). The eluted RNA (3 ul) was immediately used for one-step RT-PCR with TaqMan™ Fast Virus 1-Step Master Mix (Thermofisher, Cat# 4444436) based on the following protocol: 50°C 10 mins for reverse transcription, 95°C 20 s for RT inactivation and initial denaturation, and 48 cycles of denature (95°C 1 s) and anneal/extend (55°C 30 s).

As expected, we observed less experimental variation using Method II (single batch) than Method I (6 batches). For example, with Method I we noted a sharp drop-off of viral titers between March 26 and April 1 (Figure S1, Figure S10), which was not supported by the reprocessed single-batch data (Figure 1A, Figure 2, Figure S10). We also noted in Method I that viral titers of the northern and southern influents were nearly identical when processed together in chronological batches, but differed by a factor of 1.7 when processed as separate single batches via Method II (Figure 1B and Figure S11). Therefore, a scaling factor of 1.7 was applied to the southern influent data, generated with Method II, in order to average the two signals for several analyses (Figure 1A, 1C-F), and key findings are reproduced using the northern and southern influent data sets separately (Figure S3-S6). Our results suggest that batch effects are difficult to eliminate, even with optimized methods and internal references, and that standardization strategies (*e*.*g*., overlapping samples across batches or spiking in a surrogate virus with known concentrations) are needed to facilitate analysis of samples processed in multiple batches.

All data presented was obtained with Method II, unless specified otherwise.

### Contemporaneous societal data collection and correlation analysis

We downloaded clinical case data from March 1, 2020 to May 20, 2020 from Norfolk, Suffolk, and Middlesex Counties from Mass.gov (46). We summed the clinical cases from each county to represent the cases in the catchment of the wastewater treatment plant and calculated the new cases per day. PMMoV-corrected viral titers were averaged across replicates and influent streams. We conducted locally weighted scatterplot smoothing (LOWESS) of wastewater titers (frac=0.4) and new clinical cases (frac=0.2) purely to show the qualitative trends in Figure 1A. LOWESS smoothing was done with statsmodels.nonparametric.smoothers_lowess.lowess in python 3.6.5 and statsmodels 0.9.0. Pearson correlation was calculated between raw, unsmoothed wastewater data and raw, unsmoothed clinical data. Higher correlations were seen when comparing wastewater data to daily new clinical cases than to cumulative cases (Figure 1C and Figure S2). Moreover, cumulative cases are monotonically increasing, and do not reflect the trend of wastewater viral titers over the time, and thus we decided to do further analysis with daily new clinical cases. Correlation analysis was done in R (3.5.0).

Influenza-like illness data was downloaded from CDC FluView Portal, and includes data reported to ILINet (25, 26). Data on hospitalizations, reported deaths, number of tests administered, and daily positive test rates in Massachusetts were downloaded from Mass.gov (46). Public transit and cellular mobility data were downloaded from Massachusetts Bay Transportation Authority (30) and Citymapper (31), respectively. Supermarket visits data for Massachusetts was downloaded from SafeGraph (36).

### Social media data analysis

For analysis of emotional content, we used the dictionaries from LIWC2015 (47), which are optimized to capture social and psychological states within written text. For analysis, we focused specifically on health, anxiety, and death-related texts.

Posts from Facebook were collected by querying for posts containing the keyword “massachusetts” between January 1, 2020 and May 20, 2020 from the CrowdTangle API (35), resulting in 475,938 posts from Facebook. 105,127 (22.1%) of the posts contained words within the LIWC categories of interest.

For analysis, the available text fields from the Facebook posts were pre-processed, and psychometrics were calculated using the prevalence of words in the LIWC2015 dictionaries within the corpus of posts for a given day. Counts of words related to anxiety, death, and health were retained for analysis. The prevalence of posts about anxiety, death, and health by day was calculated by summing the counts of word matches and dividing by the number of posts in that day.

### Estimation of viral shedding function

We modeled the wastewater data W(t) as a convolution of new clinical cases *I*(*t*) and a viral shedding function into wastewater per infected individual *s*(*t*). We hypothesized that the viral shedding function could be fit by a beta distribution with parameters *α, β*, and scaling factor *c*. Since the beta distribution is defined on the range [0, 1], we sampled the probability density function for points 1/30 apart as a proxy for viral shedding over 30 days. We also hypothesized that there is some time lag in the clinical counts, which we represent as parameter *τ*. Based on the results of the correlation analysis, we back-dated the clinical cases by *τ* = 4 days to get the new cases per day function *I*(*t*). We used the numpy.convolve function to calculate the convolution of the beta distribution *s*(t) and the number of new cases per day *I*(*t*). We defined the score function as the sum of squared errors between log10(observed copies/ml in wastewater) and log10(*s*(*t*) ✲ *I*(*t*)). We used the scipy.optimize.minimize function to find parameters *α, β, c* of the beta distribution that minimized the score function. For initial parameter guesses, we used a combination of *α* = [2, 20, 50, 100, 200], *β* = [2, 20, 50, 100, 200], and *c* = [0.01, 0.1]. We multiply the optimal *s*(*t*) by an estimated wastewater volume of 1.36e6 m^3^ (1.36e12 mL) and report the total copies shed per day by each individual on both linear and log scale in Figure 1E.

To investigate the relationship between the shape of the shedding function and the clinical time lag *τ*, we conducted the optimization for *τ* ranging from 0-10 days and report the optimal shedding function for each *τ*. We did this analysis for northern and southern influent data separately, as well as the average wastewater data. We also modeled the shedding function as a gamma distribution over [0, 30] and had similar results as with the beta distribution. All shedding estimation work was done with python 3.6.5, numpy 1.14.3, pandas 0.23.0, and scipy 1.1.0.

### Poisson model simulation

We simulated wastewater titers and probability of detection under a Poisson model with equal disease incidence across the entire city. We calculated the new cases during the week of sampling by taking the difference between week 19 (May 1-7) and week 20 (May 8-14) COVID-19 case reports downloaded from the Boston Public Health Commission (48, 49). We divided by the city population and by 7 days to get the incidence rate per day (i).

For the simulation, we investigated population sizes of 100 - 40,000 people in step sizes of 100 people. We assumed each new case sheds 0.8*1.36e12 copies per day (s), as inferred in Figure 1E. Using the total wastewater volume and catchment size, we assumed each person uses 1.36e12 / 2.25e6 mL of water per day (V). For each population size n, we sampled the number of new cases (C) from a Poisson distribution with ƛ = i * n. The total viral titers for the catchment would be (C*s)/(V*n). We ran 100 trials of the simulation and plotted the simulated viral titers in Figure S8A. To calculate the probability of detection at each population size, we counted the number of trials where simulated viral titers are greater than 0. We plotted the simulation detection probability, theoretical detection probability 1-P(k=0), and observed detections in Figure S8B.

### Correlation between sewage titers, cumulative clinical rates, and demographic variables

Average sewage titers from composite samples were used for the correlation analysis. Cumulative clinical case rates (reported cases per 10,000 people) in each neighborhood were downloaded from the Boston Public Health Commission for the week of sampling and the week preceding sampling (48, 49). New cases during the week of sampling for each neighborhood were calculated by taking the difference of week 19 (May 1-7) and week 20 (May 8-14) reported cases.

Demographic data at the census block group level from 2018 were downloaded from SimplyAnalytics, Inc. (50). Each census block group is ∼1000 people, so catchments are made up of 4-30 block groups. GIS (geographic information system) data with catchment outlines was used to aggregate the demographic information for the catchment. Specifically, demographic characteristics were first assigned to each residential building within the block group. Next, resident size was estimated for each building. Finally, we calculated the population-weighted average of demographic information for each catchment.

Spearman rank correlation was calculated between sewage titers or clinical case rate and each of 38 demographic variables. p-values were adjusted using the Benjamini Hochberg method with false discovery rate of 0.1. We report all demographic variables that are significant for sewage titers and a selection of additional variables in Figure 3B. All 38 demographic variables are reported in Figure S9. Correlation analysis and multiple hypothesis correction were done with python 3.6.5, numpy 1.14.3, pandas 0.23.0, scipy 1.1.0, and statsmodels 0.9.0.

## Data Availability

The authors confirm that the data supporting the findings of this study are available within the article and its supplementary materials.

## ACKNOWLEDGMENTS

We thank the management and sampling team at the Massachusetts wastewater treatment facility who worked with us in providing the samples for analysis, in particular Conor Donovan, Jim Fitzgerald, Louis Logan, Nicole Mangano, Keith Stocks, Sean Winter, and David Wu. We thank Lisa Wong (MWRA) for providing flow data, and Stephen Estes-Smargiassi (MWRA) and Betsy Reilley (MWRA) for helpful discussion. We thank Penny Chisholm (MIT) and Allison Coe (MIT) for access to equipment and other supplies, Mathilde Poyet (MIT) and Shandrina Burns (MIT) for logistical support, Andrew Tang (Broad Institute) for assistance with Figure 2, Nicholas Santos-Powell for thoughtful comments on the manuscript, Kyle Bibby (University of Notre Dame) for suggestions on PMMoV, and Karina Gin (National University of Singapore) and Lee Ching Ng (National Environmental Agency, Singapore) for helpful discussion and for sharing the Amicon protocol. We also thank Dr. Victor M. Corman (Charité Universitätsmedizin, Germany) for sharing the data of viral concentrations in patients’ stool samples, and the SafeGraph team for providing free access to their data for COVID-19-related research. Finally, we express our deep gratitude to all healthcare professionals and first-line responders who have been caring for patients with COVID-19.

This work was supported by the Center for Microbiome Informatics and Therapeutics and Intra-CREATE Thematic Grant (Cities) grant NRF2019-THE001-0003a to JT and EJA; National Institute on Drug Abuse of the National Institutes of Health award numbers K23DA044874; and R44DA051106 to MM and PRC, Hans and Mavis Psychosocial Foundation funding, and e-ink corporation funding to PRC; funding from the Morris-Singer Foundation and NIH award R01AI106786 to WPH; funds from the Massachusetts Consortium on Pathogen Readiness and China Evergrande Group to TBE, PRC, MM, and EJA; funding from the Singapore Ministry of Education and National Research Foundation through an RCE award to Singapore Centre for Environmental Life Sciences Engineering (SCELSE) to SW and JT; and a National Institute of General Medical Sciences of the National Institutes of Health award, number R01GM130668, to MS. The content is solely the responsibility of the authors and does not necessarily represent the official views of the funding institutions.

---

**Figure S1.**
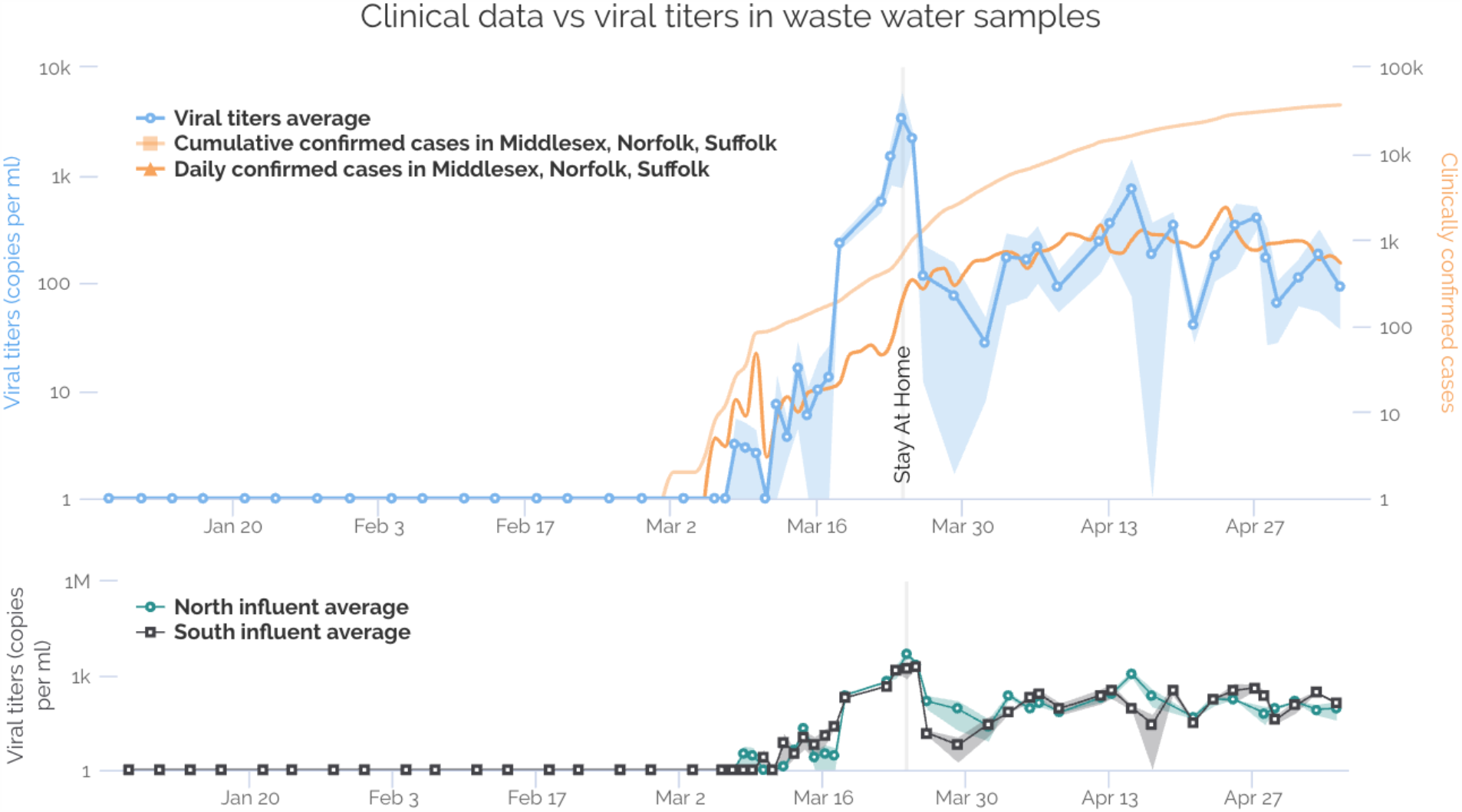
Viral titers in wastewater samples processed in chronological groups with Method I. 108 sewage samples (January 8 to May 5) were collected from northern and southern catchments. Top: Viral titers in the wastewater (blue line along the primary y-axis), daily (orange line) and cumulative (light orange) confirmed cases in MA along the secondary y-axis. Viral titers are calculated and averaged from qPCR results with N1 and N2 primers (normalized with PMMoV reference). Shaded area represents the minimum and maximum values. The earliest dates of SARS-CoV-2 detection in sewage samples were March 8 for Northern influent and March 10 for Southern influent. Bottom: Viral titers in northern and southern influents. Grey vertical line: first day of stay-at-home order.

**Figure S2.**
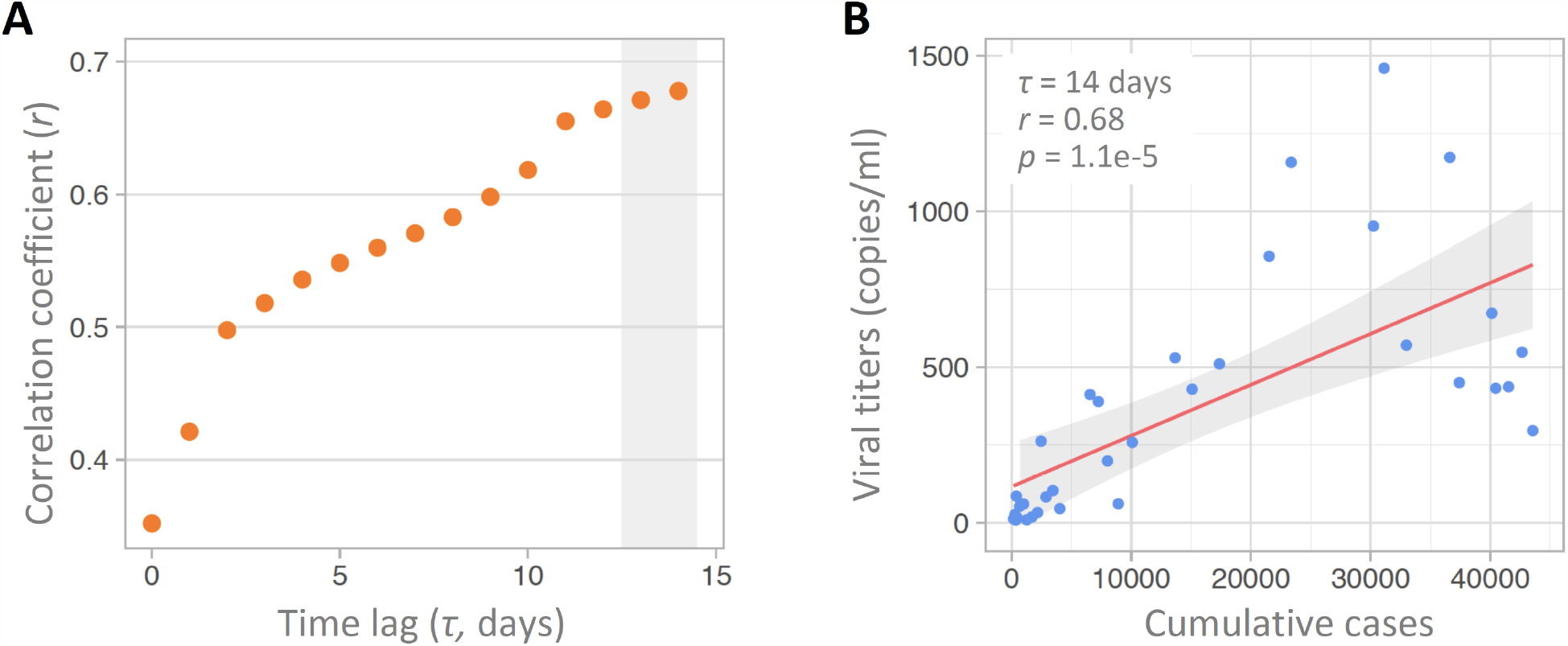
Correlation analysis between viral titers in wastewater and clinically reported cumulative cases, with or without a time lag. (A-B) Linear correlation between unsmoothed viral titers in the northern and southern influents and the number of cumulative cases with different time lags from 0 to 14 days. (B) Viral titers correlate with cumulative cases with a 14 day time lag. Red solid line is the linear regression fitting. Grey area: 95% confidence interval from standard error of the fitting. Pearson’s r = 0.68, p = 1.1e-5.

**Figure S3.**
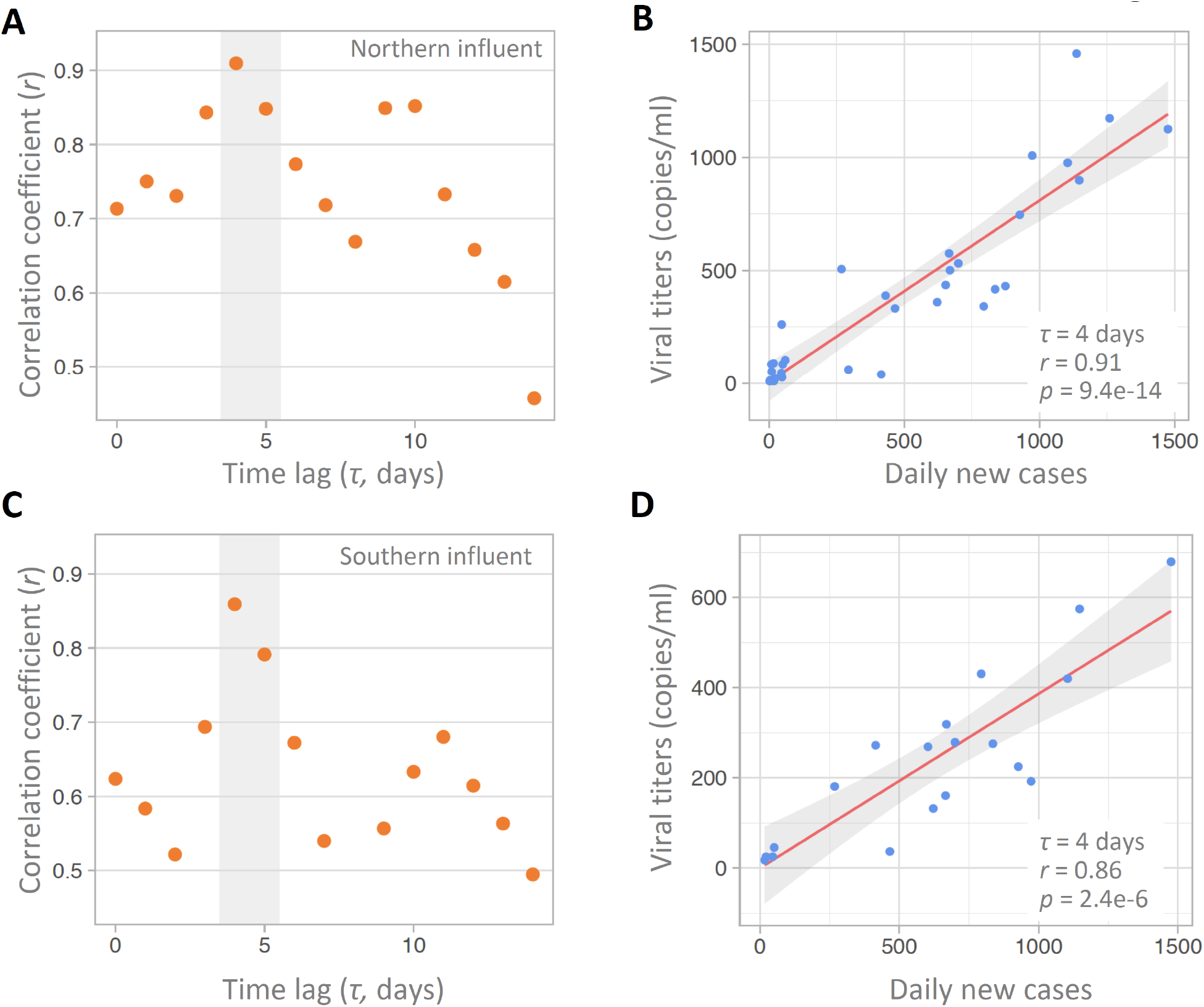
Correlation analysis between viral titers in northern or southern influents and daily new cases, with or without a time lag. (A-B) Linear correlation between unsmoothed viral titers in northern influent and unsmoothed daily new cases with different time lags from 0 to 14 days. Pearson correlation coefficient is highest with a 4 day time lag, with Pearson’s r = 0.91 and p = 9.4e-14. (C-D) Linear correlation between unsmoothed viral titers in southern influent and unsmoothed daily new cases with different time lags from 0 to 14 days. Pearson correlation coefficient is highest with a 4 day time lag, with Pearson’s r = 0.86 and p = 2.4e-6. Red solid line is the linear regression fitting. Grey area: 95% confidence interval from standard error of the fitting.

**Figure S4.**
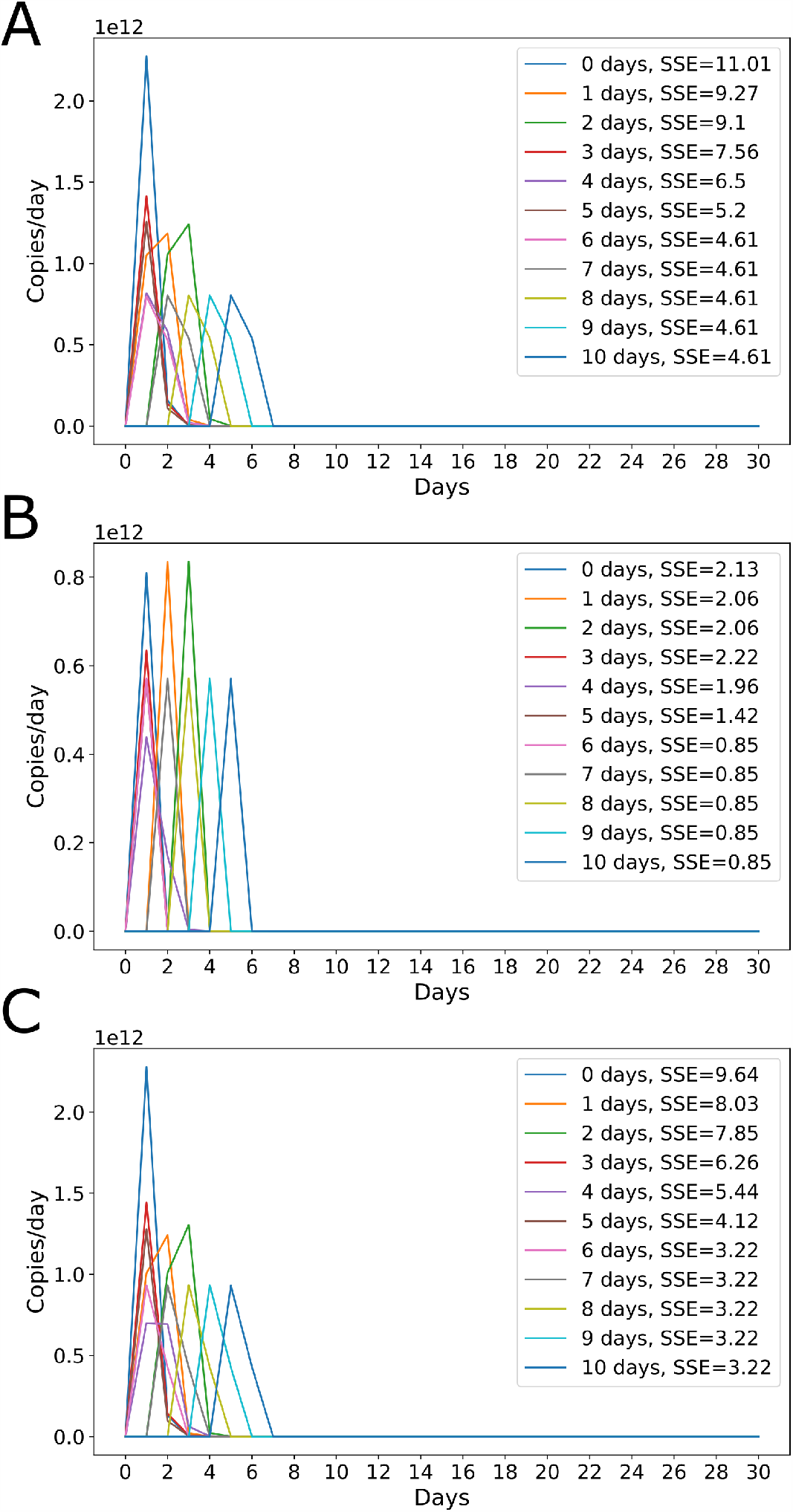
Optimal viral shedding function for various time lags. Allowing clinical time lag to vary between 0-10 days reveals optimal shedding functions for (A) northern influent, (B) southern influent, (C) average of southern and northern influent data sets. The optimal shedding function is sharply peaked for all time lags. The optimal shedding function was multiplied by average wastewater volume of 1.36e6 m^3^ (1.36e12 mL) to convert to copies shed per day.

**Figure S5.**
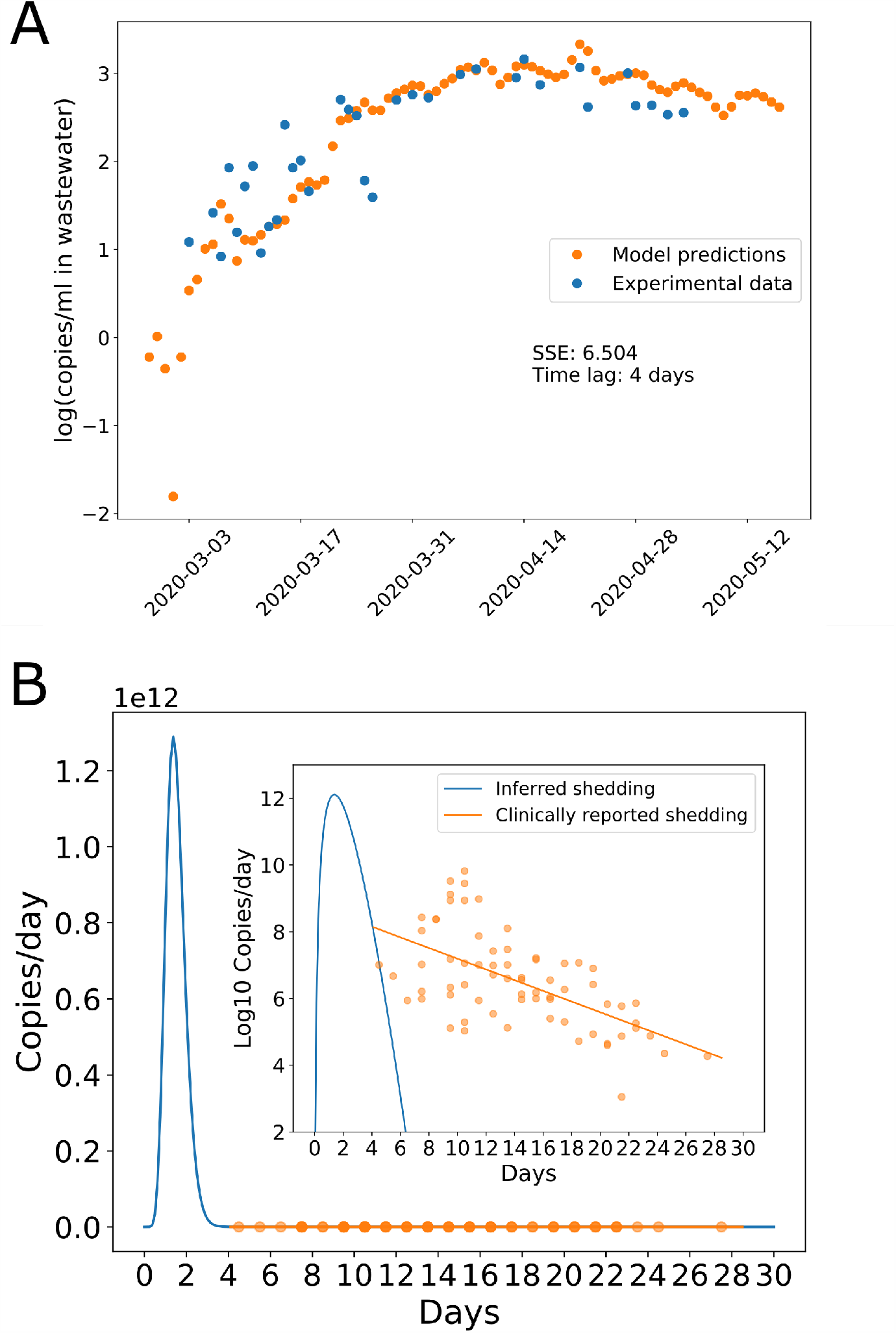
Model fit and optimal shedding function for northern influent alone, with 4 day time lag. (A) The optimal shedding function minimizes the sum of squared errors between wastewater data (log10) and the convolution of new cases per day *I*(*t*) with the individual shedding function *s*(*t*) (B). Clinically reported shedding data are shown in (B) for reference, with linear regression fit (18). Inset shows shedding on a log scale.

**Figure S6.**
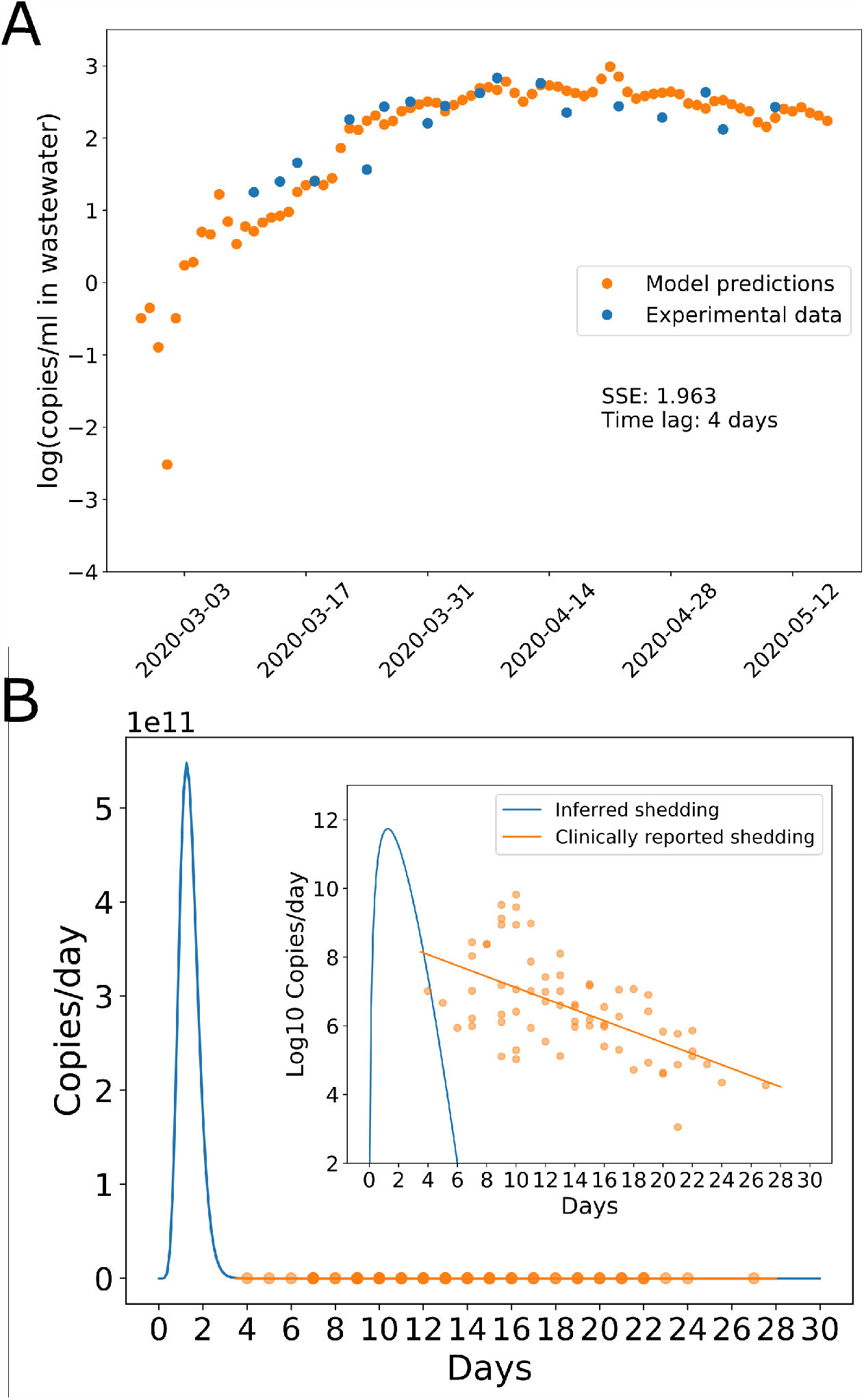
Model fit and optimal shedding function for southern influent alone, with 4 day time lag. (A) The optimal shedding function minimizes the sum of squared errors between wastewater data (log10) and the convolution of new cases per day *I*(*t*) with the individual shedding function *s*(*t*) (B). Clinically reported shedding data are shown in (B) for reference, with linear regression fit (18). Inset shows shedding on a log scale.

**Figure S7.**
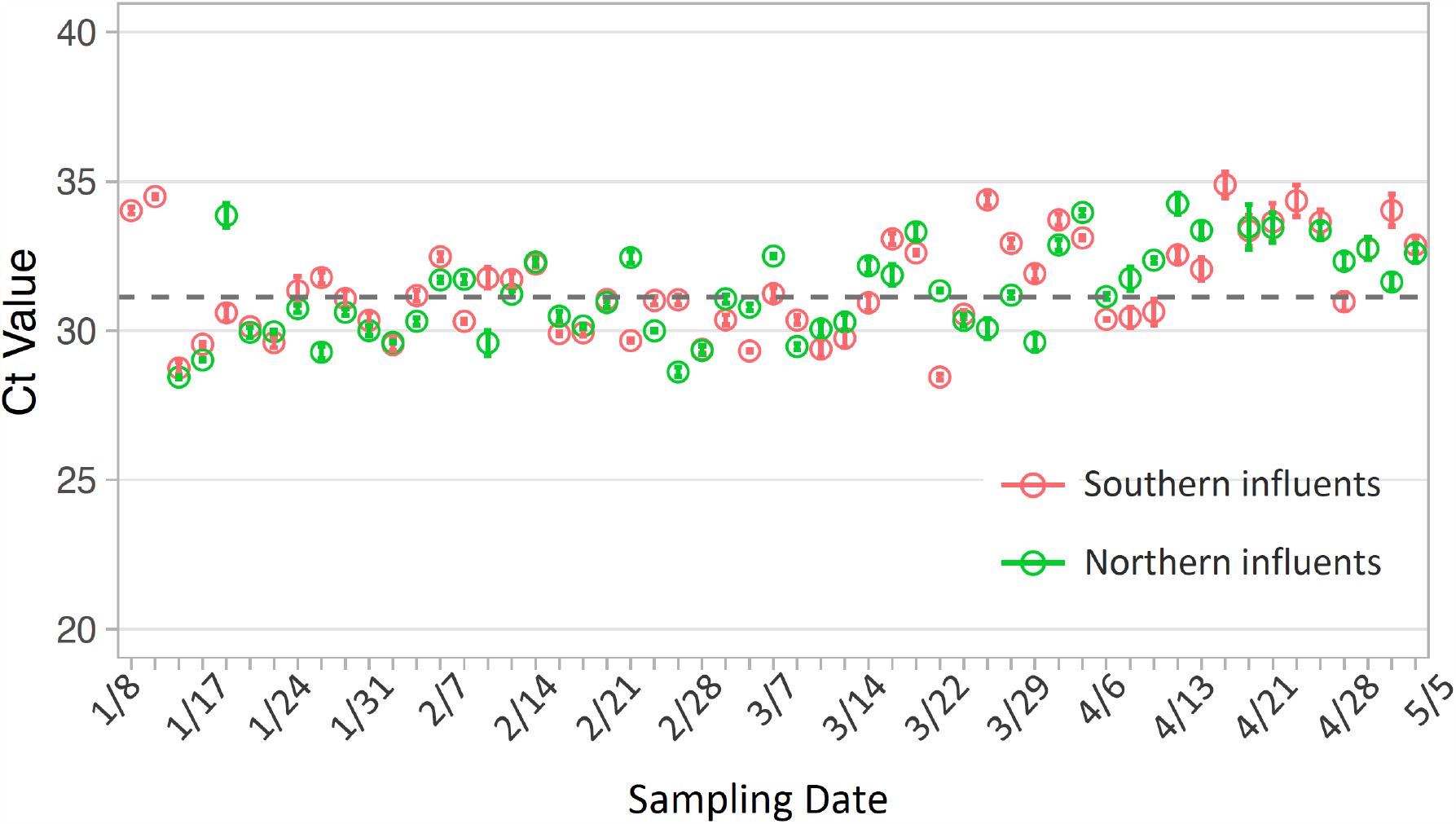
PMMoV Ct values in all the sewage samples collected from January to early May. PMMoV in northern and southern influents varies little over time with the exception of late March. Grey dashed line: median of the Ct value in all the samples. Each data point is the mean ± sd (standard deviation) from three replicates.

**Figure S8.**
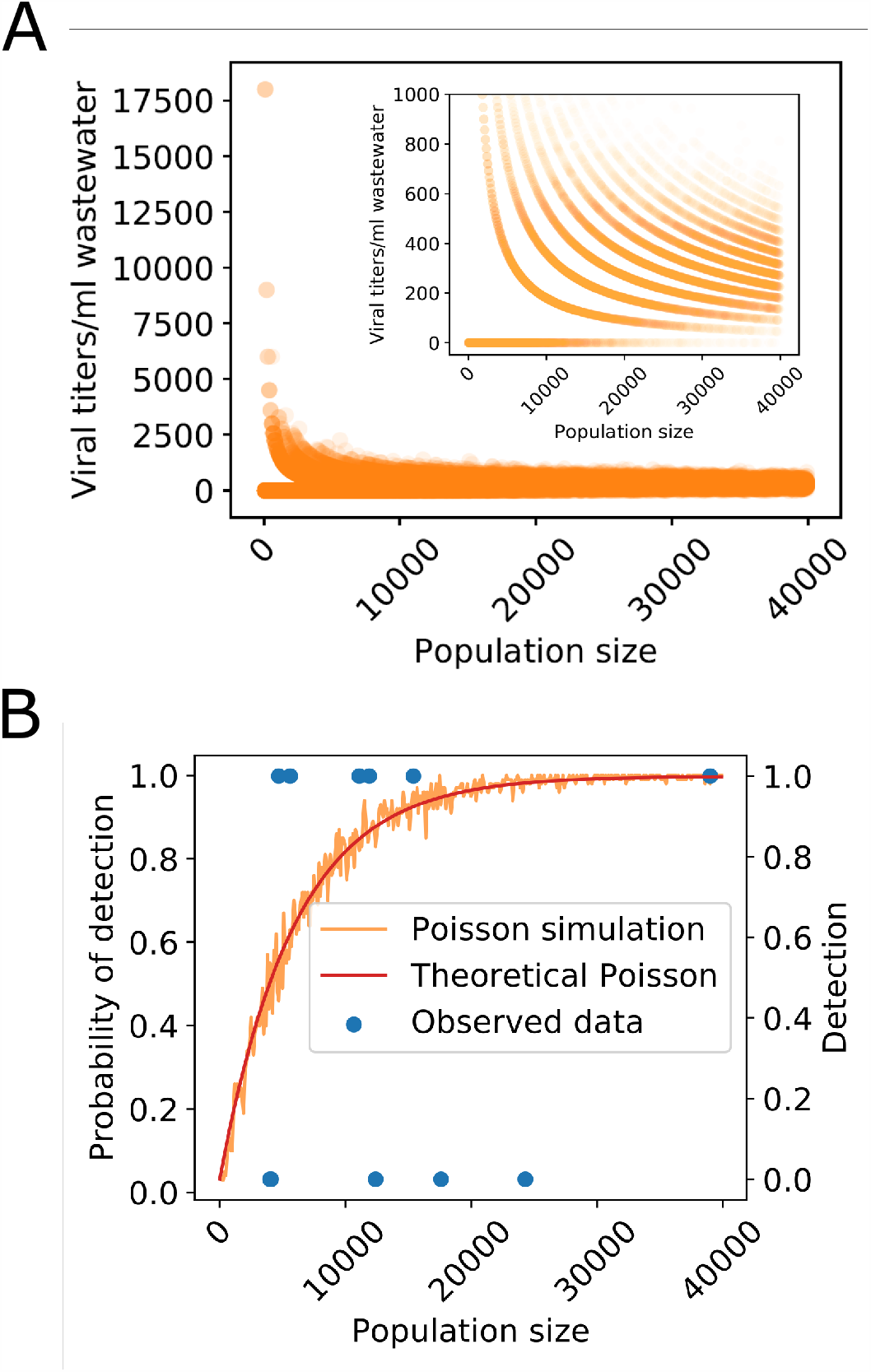
Observed SARS-CoV-2 detection at Massachusetts catchments strongly deviates from a Poisson model that assumes equal incidence across all neighborhoods. (A) Simulating viral titers in different sized catchments using an incidence of 0.00017 (new cases in city during week of sampling / city population / 7 days) and the peak shedding inferred in Figure 1E. Simulated viral titers have high variance at low population sizes, with many non-detects and a few extreme titers. Viral titers converge to the average as population size increases. Inset: zoom in on 0-500 copies/ml wastewater. (B) Observed SARS-CoV-2 detection compared to a theoretical Poisson model with uniform incidence = 0.00017 and the Poisson simulation in (A). Catchments with populations > 15000 should have 100% detection based on the Poisson null model, but observed wastewater detection is 0 for some of those catchments.

**Figure S9.**
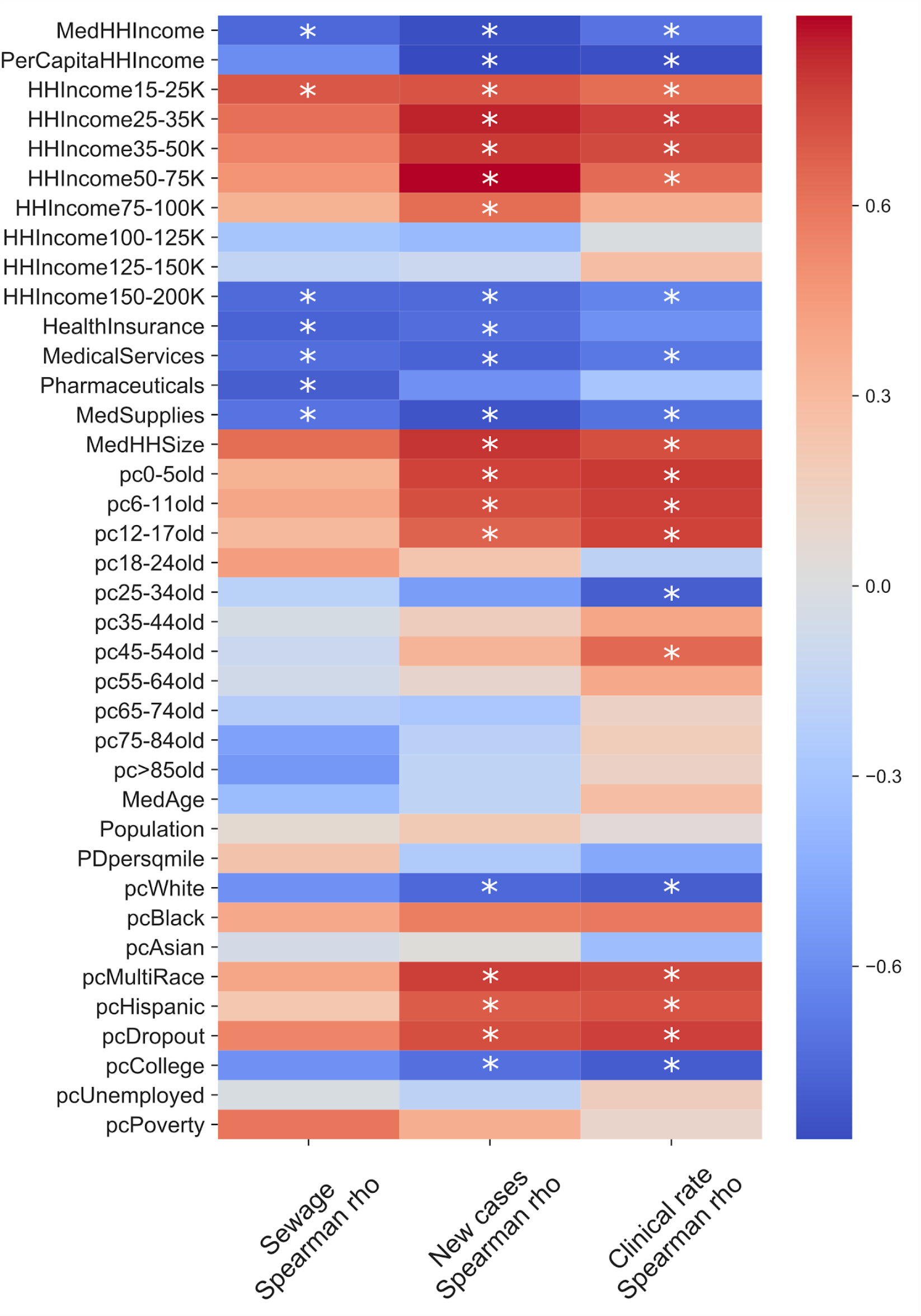
Spearman correlations between sewage titers/clinical data and demographic variables in 11 Boston catchments reveal stronger correlations with demographic variables than with population size. Asterisk denotes significant p-value at FDR < 0.1. pc: percent, HH: household, Ave: average, Med: median. MedHHIncome, PerCapitaHHIncome: Median and Per capita household income. HHIncome15-25K: percentage of population with household income within the range 15-25K throughout the year. HealthInsurance, MedicalServices, Pharmaceuticals, MedSupplies: average household expenditure on health insurance, medical services, pharmaceuticals, and medical supplies. MedHHSize: Median household size. pc0-5old: percentage of individuals aged 0-5 years. MedAge: Median age. Population: catchment population size. PDpersqmile: population density per square mile. pcWhite: percentage of individuals identifying as white. pcDropout: percentage of individuals who dropped out of high school. pcCollege: percentage of individuals who completed college. pcUnemployed: percentage of people who are unemployed. pcPoverty: percentage of population in poverty.

**Figure S10.**
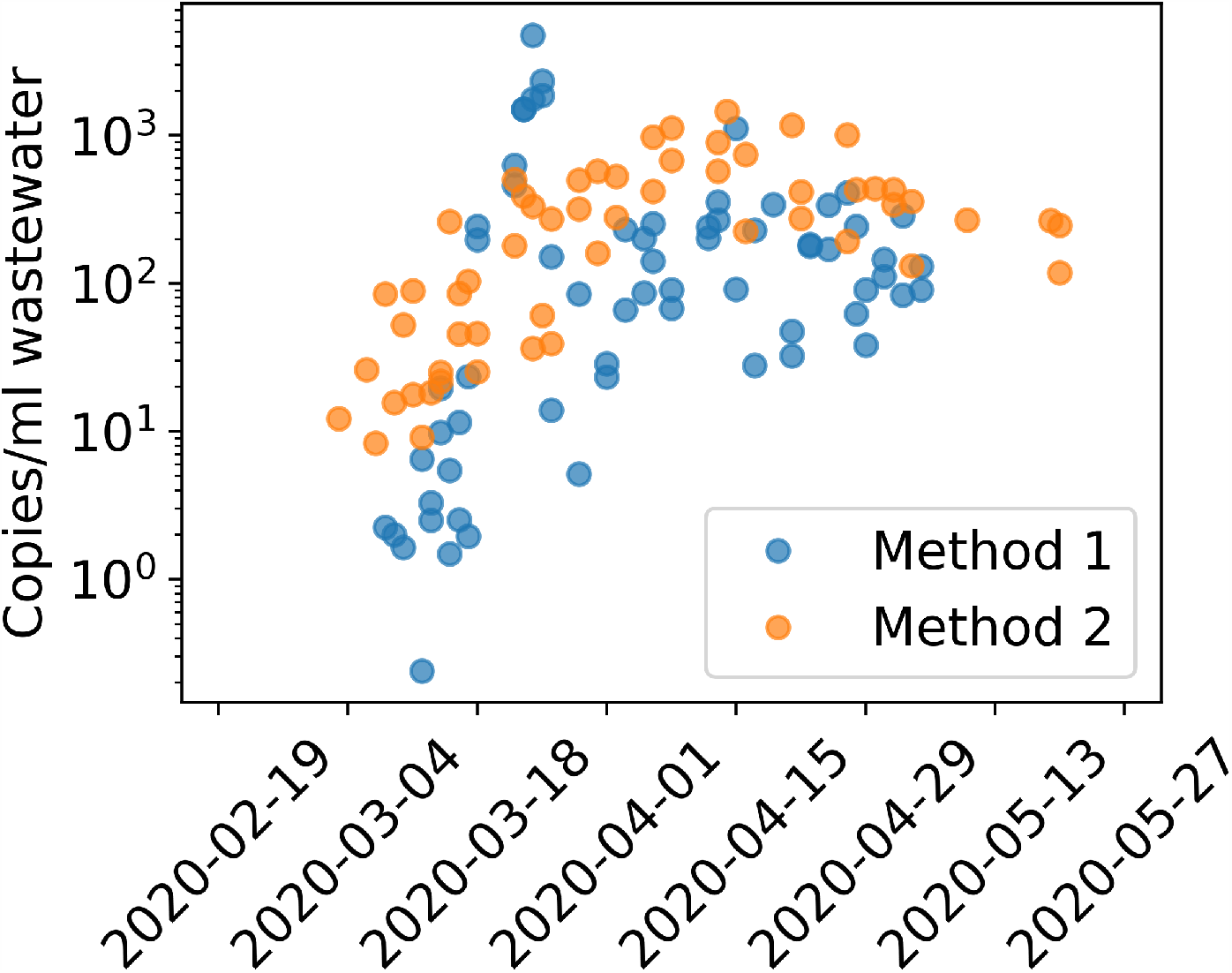
Average normalized viral titers for northern and southern influents using Method 1 vs Method 2 of viral particle precipitation, RNA extraction, and RT-qPCR. Method 1 has a sharp peak and drop off for samples processed in the 3/18-3/25 batch, whereas Method 2 has a smooth increase and plateau.

**Figure S11.**
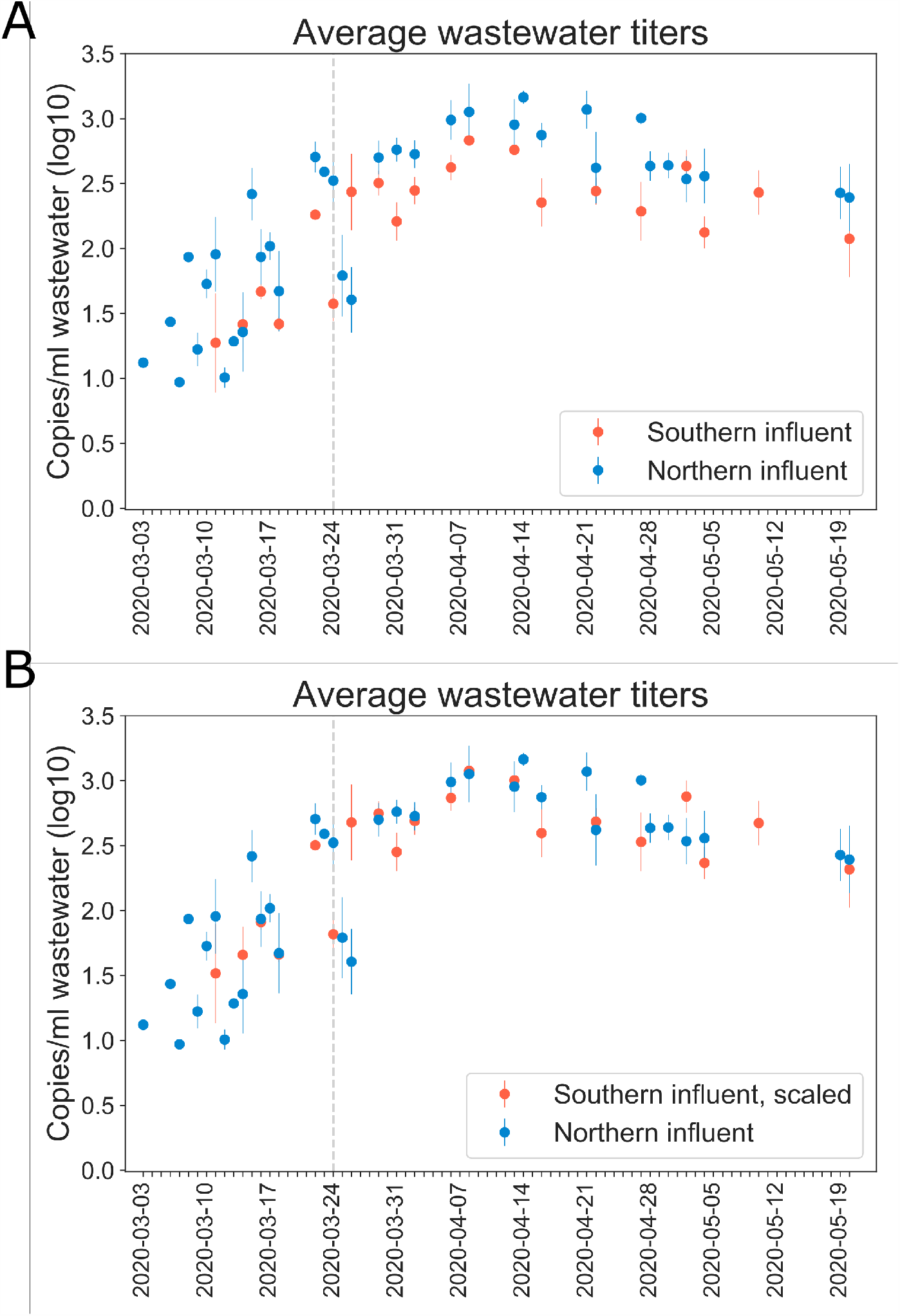
Viral titers in wastewater samples processed with Method II. Samples in northern and southern influents were each processed in one batch. (A) The trends of viral titers in northern and southern influents agreed well, however, absolute value of viral titers differed by a factor of 1.7 that was not observed in Method I. (B) Scaled southern influent data is plotted to match northern data magnitudes. The scaling factor *k* was found by minimizing the sum of squared errors between southern data * *k* and northern data. Grey vertical line: first day of stay-at-home order in Massachusetts.

